# Trying to Find the Answer for Two Questions in Patients with COVID-19: *1. Is pulmonary infiltrate of COVID-19 infective or inflammatory in nature (Pneumonia or Pneumonitis)? 2. Is Hydroxychloroquine plus Azithromycin or Favipiravir plus Dexamethasone more effective in the COVID-19 treatment?*

**DOI:** 10.1101/2020.08.25.20181388

**Authors:** Adem Dirican, Tugce Uzar, Irem Karaman, Aziz Uluisik, Sevket Ozkaya

## Abstract

**Background:** During the current pandemic, a great effort is made to understand the COVID-19 and find an effective treatment. As of 17 August 2020, there is no specific drug or biologic agent which have been approved by the FDA for the prevention or treatment of COVID-19.

**Methods:** We retrospectively analyzed the clinical and radiological findings of 211 COVID-19 in-patients that were treated between March - August 2020. Confirmation of a COVID-19 diagnosis was made according to a positive RT-PCR result with a consistent high-resolution-CT (HRCT) finding. Radiological images and the rate of clinical response of patients were investigated.

**Result:** While 128 patients (58.7) did not develop pneumonia, the mild, moderate and severe pneumonia ratios were 28(13.2%), 31(18.7%) and 27(22.9%). 72 patients (34.1%) whose PCR tests were positive did not show any symptom and they were followed in isolation without treatment. 52 patients (24.6%) received hydroxychloroquine plus azithromycin, 57 patients (27%) received favipiravir and 30 patients (14.2%) received favipiravir plus dexamethasone as the first line of treatment. 63.1% of pneumonia patients who received hydroxychloroquine plus azithyromycine, 28.3% of patients who received favipiravir and 10% of patients who received favipiravir plus dexamethasone showed a failure of treatment.

**Conclusion:** The pulmonary infiltrates of COVID-19 are not infective; therefore, the characteristic of the disease should be described as COVID-19 pneumonitis instead of pneumonia. The favipiravir plus dexamethasone seems to be the only drug combination to achieve the improvement of radiological presentation and clinical symptoms in COVID-19 pneumonia patients.

## Introduction

After the coronavirus disease 2019 (COVID-19) emerged in the Wuhan, China in late 2019 from a zoonotic source; the outbreak spread rapidly throughout the world. On March 11 2020, World Health Organization (WHO) declared the COVID-19 outbreak a pandemic (1). Although the majority of COVID-19 cases are either asymptomatic or mild, COVID-19 may cause a respiratory compromise requiring hospitalization and supportive care in a significant proportion of patients (2). As of August 17 2020, this pandemic caused the death of more than 772.000 people with a significant economical and sociological burden globally (3). Besides its high infectivity rate, COVID-19 also has high morbidity and mortality due to autoimmune destruction of the lungs stemming from the release of the pro-inflammatory cytokines (2).

It is known that; the severe acute respiratory syndrome coronavirus 2 (SARS-CoV-2) mainly affects the respiratory tract and reaches the lungs, eventually causes the potentially fatal pneumonia in severe cases (2). Since the studies investigating the pathophysiological characteristic of this novel disease are still ongoing, the search for effective therapies requires a comprehensive approach. As of 17 August 2020, there is no specific drug or biologic agent which is approved by the FDA for the prevention or treatment of COVID-19 (4). Numerous other antiviral agents, immunotherapies, and vaccines continue to be investigated and used as potential therapies in the clinical trials. Several guidelines and reviews of pharmacotherapy for COVID-19 investigating the potential drugs have also been published (4,5). Existing drugs with well-documented effects including hydroxychloroquine, azithromycin, favipiravir and dexamethasone are being used as a combination or alone to diminish the symptoms of respiratorily compromised severe COVID-19 patients. (4,5).

To find an effective treatment, the most important point to be illuminated in patients with COVID-19; is the question of whether lesions in the lung are infective or inflammatory in nature. Therefore, in this retrospective study, we aimed to find the answer of the two questions: “Is pulmonary infiltrate of COVID-19 infective or inflammatory in nature?” and “Is Hydroxychloroquine plus Azithromycin or Favipiravir plus Dexamethasone more effective in the COVID-19 treatment?”.

## Material & Method

### Study design

We retrospectively analyzed the radiological and clinical findings of COVID-19 in-patients and out-patients that were treated in Samsun VM Medicalpark Hospital between March-August 2020. This study was approved by the Institutional Review Board of SB (IRB No. 2020-07-21T10-25-53) and performed in accordance with the principles of the Declaration of Helsinki.

### Patient Selection

Patients who were suspected to be infected with COVID-19 were confirmed with laboratory results and included in the study. Only laboratory-confirmed cases were included in the study. A total of 211 patient has been included in the study. All the recovered patients were discharged with 2 consecutive negative RNA detection (interval above 24 h) and clinical improvement and followed for another 14 days after discharge.

### Patient Grouping

Patients with pneumonia were divided into three groups by means of severity as mild, moderate and severe depending on the total volume of radiologic infiltrations. Small and by one by ground-glass opacities or consolidations were considered as mild pneumonia (Figure 1). Those with a total volume of diffuse ground-glass opacities or consolidations, smaller than the volume of right upper lobe were considered as moderate pneumonia (Figure 2). Severe pneumonia was defined as the total volume of diffuse ground-glass opacities bigger than the volume of right upper lobe (Figure 3).

**Figure 1.**
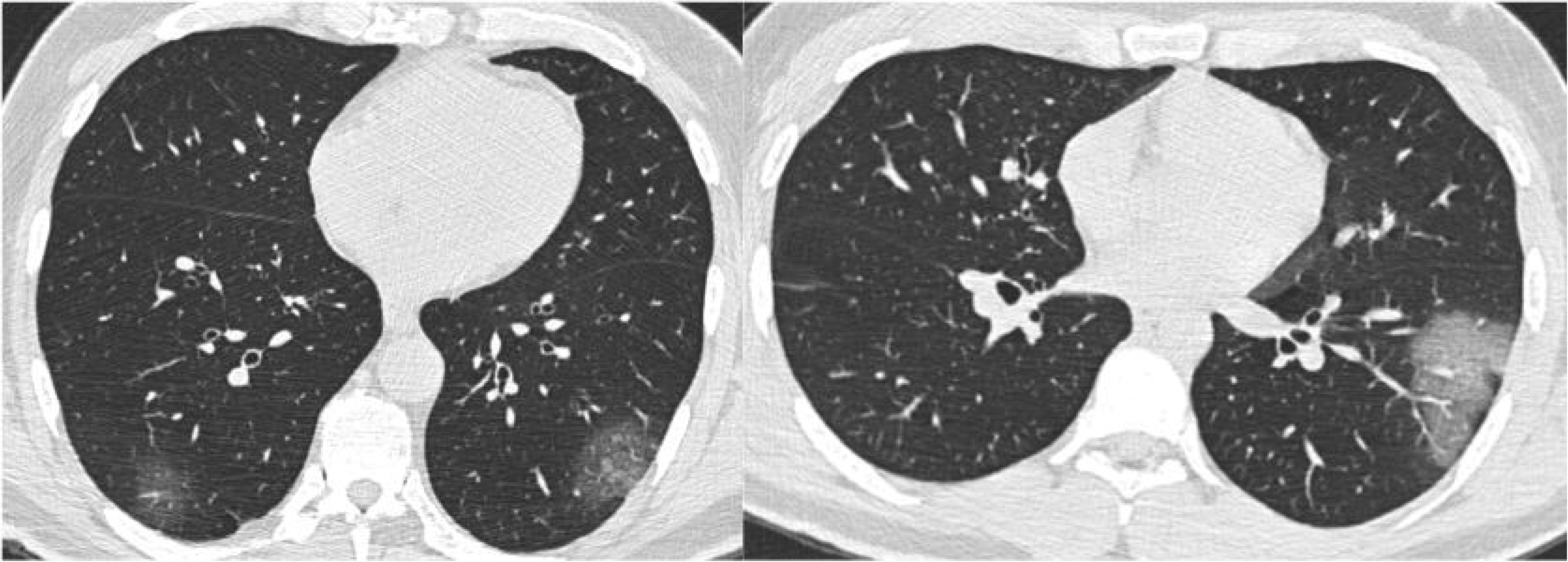
70 and 30 years-old two men admitted to hospital with complaints of cough and fever. Their COVID-19 PCR test results were positive. Their HRCT demonstrated a mild case of COVID 19 pneumonia with ground glass infiltrates.

**Figure 2.**
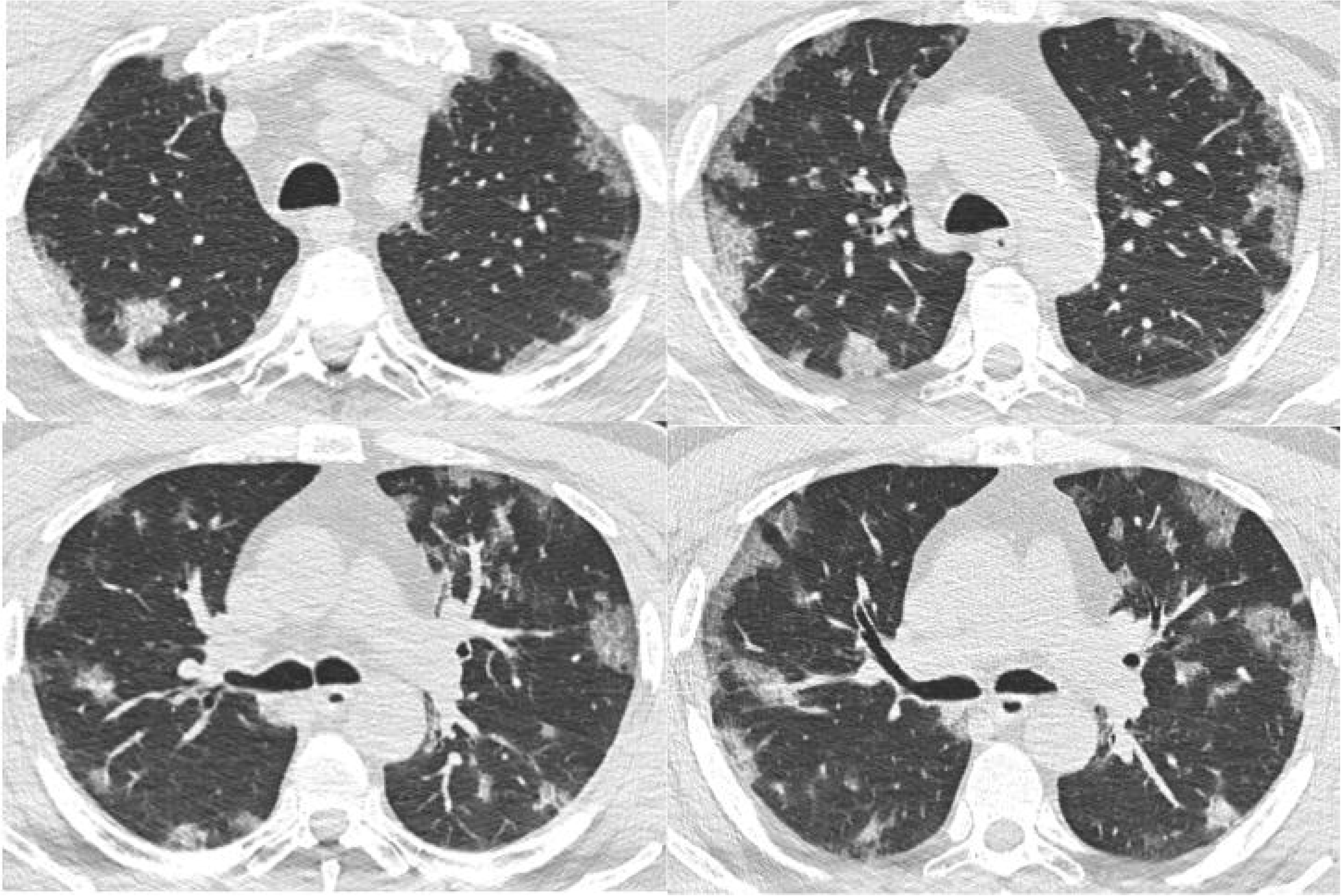
A 54-year-old male patient admitted to hospital with complaints of cough and dyspnea. His COVID-19 PCR test was positive. His HRCT showed that he had COVID-19 moderate pneumonia with bilateral, subpleural and peripheral ground glass opacities (GGOs) and crazy paving appearance (GGOs and inter-/intra-lobular septal thickening).

**Figure 3.**
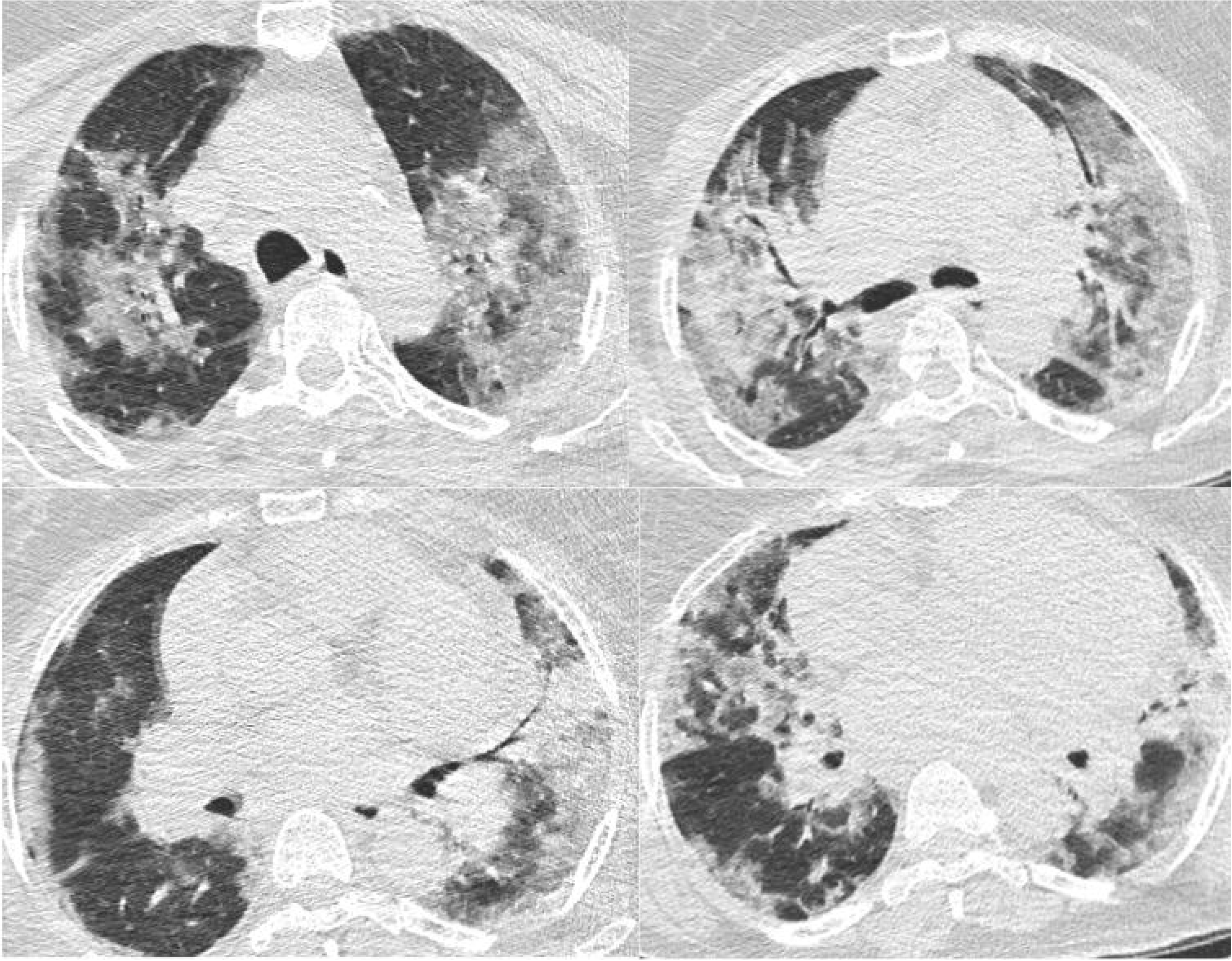
A 68-year-old female patient referred to ICU because of respiratory failure. Her COVID-19 PCR test was positive. Her HRCT demonstrated a severe case of COVID-19 pneumonia with bilateral, diffuse air space consolidations and bilateral, subpleural and peripheral ground glass opacities (GGOs).

### COVID-19 testing

Respiratory samples were obtained by nasopharyngeal and nasal swabs and analyzed by reverse-transcriptase-polymerase-chain-reaction (RT-PCR) assay. Confirmation of a COVID-19 diagnosis was made according to a positive RT-PCR result with a consistent high-resolution-CT (HRCT) finding. Also, the patients whose multiple nasopharyngeal swab samples were negative for SARS-CoV-2 but the serological IgM/IgG antibody was positive in the rapid antibody test included in the study.

### Treatment Protocol

In the beginning, all patients were prescribed with oral hydroxychloroquine sulfate 400 mg two times in the first day as the loading dose and, 200 mg two times per day for the following four days. Azithromycin 500 mg was prescribed as the loading dose on the first day and 250 mg one time per day for the following four days. Favipiravir was loaded as 1600 mg two times in the first day as the loading dose and 600 mg two times for the following four days as the maintenance dose. The dose of the systemic steroid (dexamethasone or methylprednisolone) was adjusted according to daily status of the patient. All the patients who seem to recover by fibrosis were prescribed as long-term low dose oral steroid. The treatment was considered as effective if clinical and radiological recovery was observed at the end of the treatment.

### Statistical Analysis

The quantitative data were described as the mean ± standard deviation, or as the median (min - max). The qualitative data were described by the number of cases (proportion, %). All of the analysis was performed using SPSS Version 22.0.

## Results

A total number of 211 patient was included in the study. The characteristics of the patients were shown in Table 1. The nasopharyngeal swab PCR tests were positive in 97.2% of patients and serological IgM/IgG antibody against SARS-CoV-2 was positive in 2.8% of patients at the time of diagnosis. The mean age was 63.2 years old and male to female ratio was 110/101. The number of pregnant, <18 years old and >65 years old patients were 7(3.3%), 23(10.9%), and 36(17%), respectively. 81 patients (38.3%) had pulmonary infiltrates on HRCT. There was no pneumonia in pregnant patients and patients who were <18 years old. 10 patients (4.7%) developed respiratory failure and 3 patients (30% in ICU and 1.4% of total patients) died during the follow-up time.

**Table I.** The Characteristics of Patients.

|  | **n(%)** |
| --- | --- |
| *Age (mean ±SD)* | 61.7 |
| **Gender** |  |
| *Male/Female* | 110/101 |
| **Symptoms (n,%)** |  |
| *Cough* | 183(86.7) |
| *Fever* | 125(59.2) |
| *Dyspnea* | 64(30.3) |
| **Pregnant** | 7(3.3) |
| **Health Worker** | 18(8.5) |
| **<18 years old** | 23(10.9) |
| **>65 years old** | 36(17) |
| **Diagnostic Test** |  |
| *PCR\** | 205(97.1) |
| *Antibody (IgG/Ig M)* | 6(2.8) |

\*PCR: Polymerase-Chain-Reaction

The mild, moderate and severe pneumonia ratios were 28(13.2%), 31(18.7%) and 27(22.9%), respectively as shown in Table II. The radiologic images of each group of patients were shown in Figures 1,2 and 3.

**Table II.** COVID-19 Pneumonia Status of the Patients.

| **Pneumonia** | **n(%)** |
| --- | --- |
| None | 124(58.7) |
| Mild | 28(13.2) |
| Moderate | 31(14.6) |
| Severe | 27(12.7) |

The given drugs and treatment failure rates were shown in Table III. 72 patients (34.1%) whose PCR tests were positive did not show any symptom. They did not receive any treatment and were observed in the isolation for 14 days. 52 patients (24.6%) received hydroxychloroquine plus azithromycin, 57 patients (27%) received favipiravir and 30 patients (14.2%) received favipiravir plus dexamethasone in the first place. 63.1% of pneumonia patients who received hydroxychloroquine plus azithromycin showed a failure of treatment as shown in Figure 4A-4B, 5A-5B, and 6A-6B. The patients that have treatment failure were successfully re-treated with favipiravir plus dexamethasone and their radiological infiltrates were shown in Figures 4C, 5C and 6C.

**Table III.** Treatment Results.

|  | **n(%)** |
| --- | --- |
| **Pneumonia** | 87(41.2) |
| **ICU\*** | 12(5.6) |
| **Mortality** | 3(1.4) |
| **Treatment** |  |
| *HQ\*\*+Azithromycin* | 52(24.6) |
| *with pneumonia* | 38(18) |
| *treatment failure* | 24(63.1) |
| *Favipiravir* | 57(27) |
| *with pneumonia* | 53(25.11) |
| *treatment failure* | 15(28.3) |
| *Favipiravir+CS\*\** | 30(14.2) |
| *with pneumonia* | 30(14.2) |
| *treatment failure* | 3(10) |

\***ICU:** Intensive Care Unit,
\*\***HQ:** Hydroxychloroquine
\*\*\***CS:** Corticosteroid

**Figure 4A.**
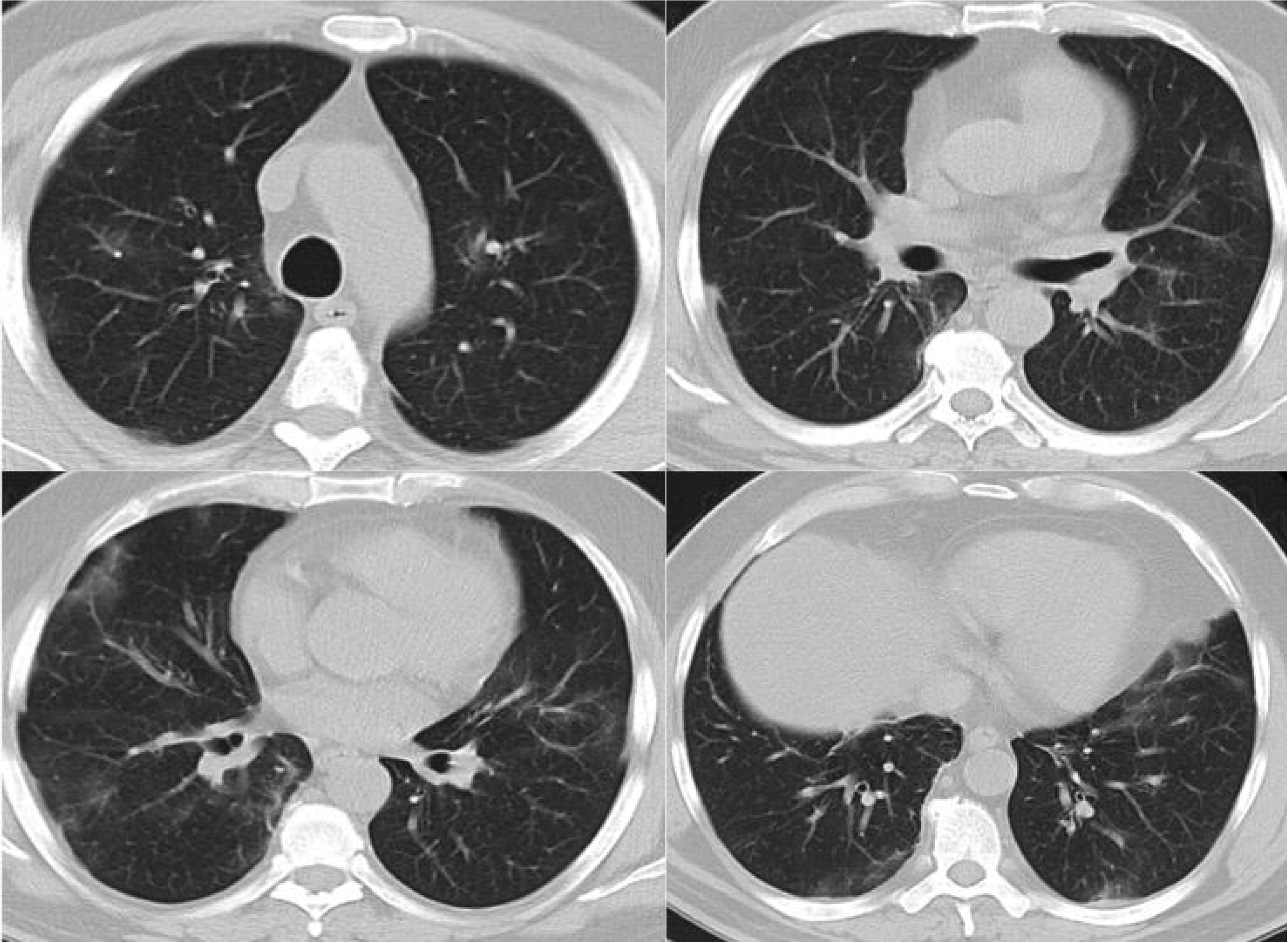
A 57-year-old male patient presented with the complaints of cough and fever. His COVID-19 PCR test was positive. Due to the ground glass infiltrates that were seen at thorax HRCT, treatment with hydroxychloroquine plus azithromycin was started.

**Figure 4B.**
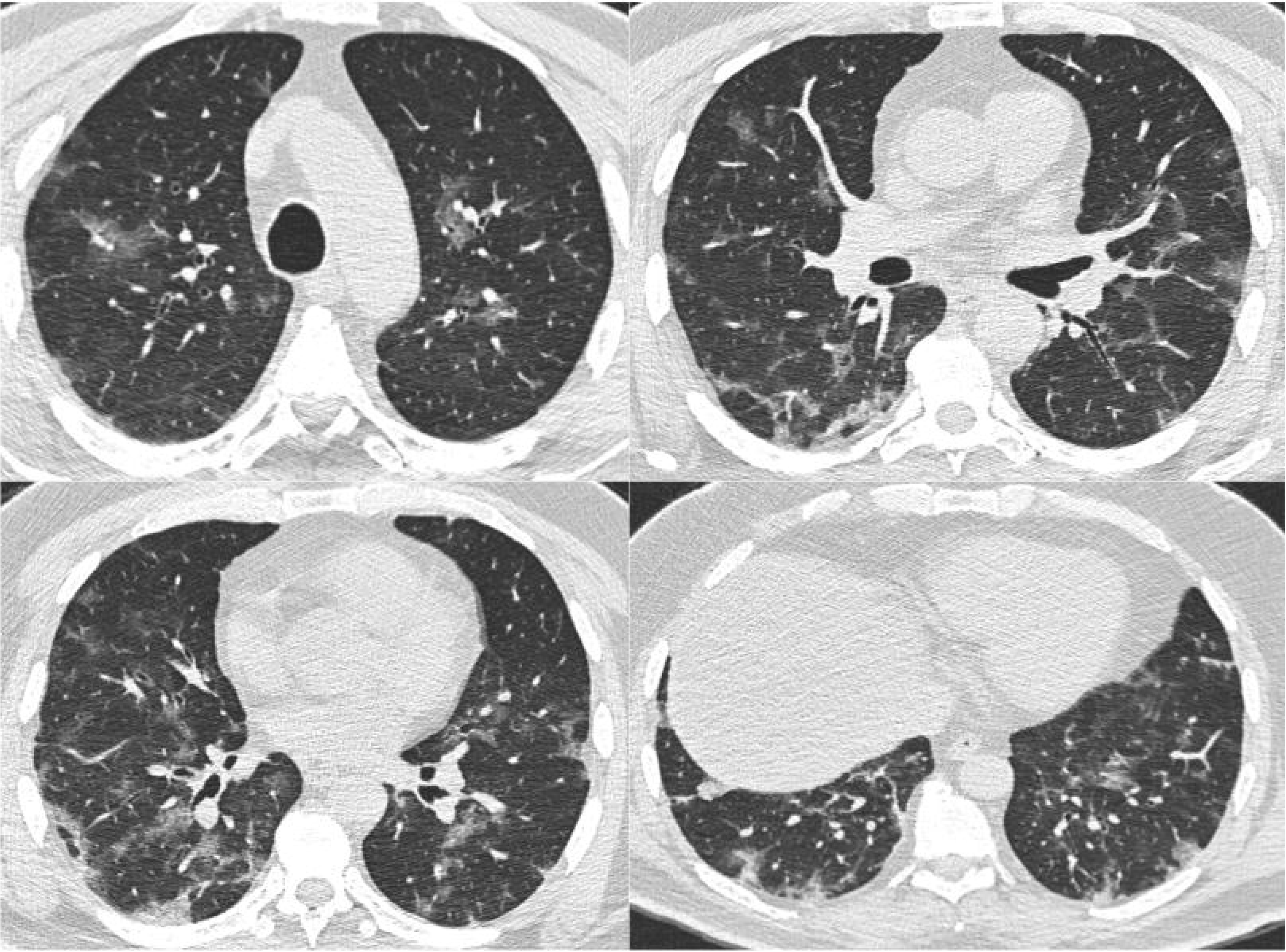
Despite five days of treatment with hydroxychloroquine plus azithromycin, the fever and cough complaints of patient in Figure 4A did not resolve. Since the control CT on the 7th day revealed an increase in the ground-glass infiltrates, treatment with favipiravir plus dexamethasone was started.

**Figure 4C.**
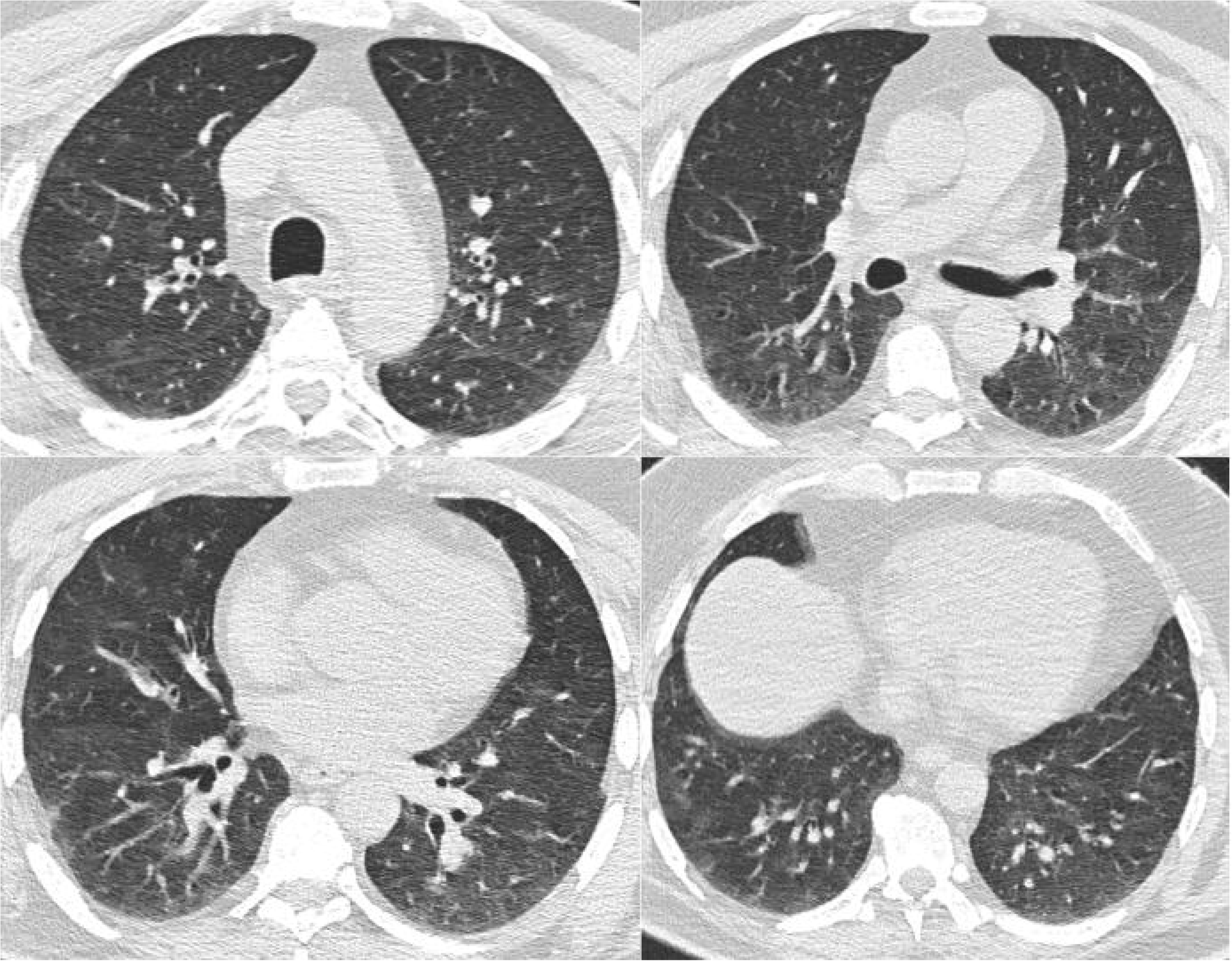
After five days of treatment with favipiravir plus dexamethasone, chest x-ray on the 7^th^ day revealed the regression of lesions and patient’s complaints (cough and fever) have shown a significant improvement.

**Figure 5A.**
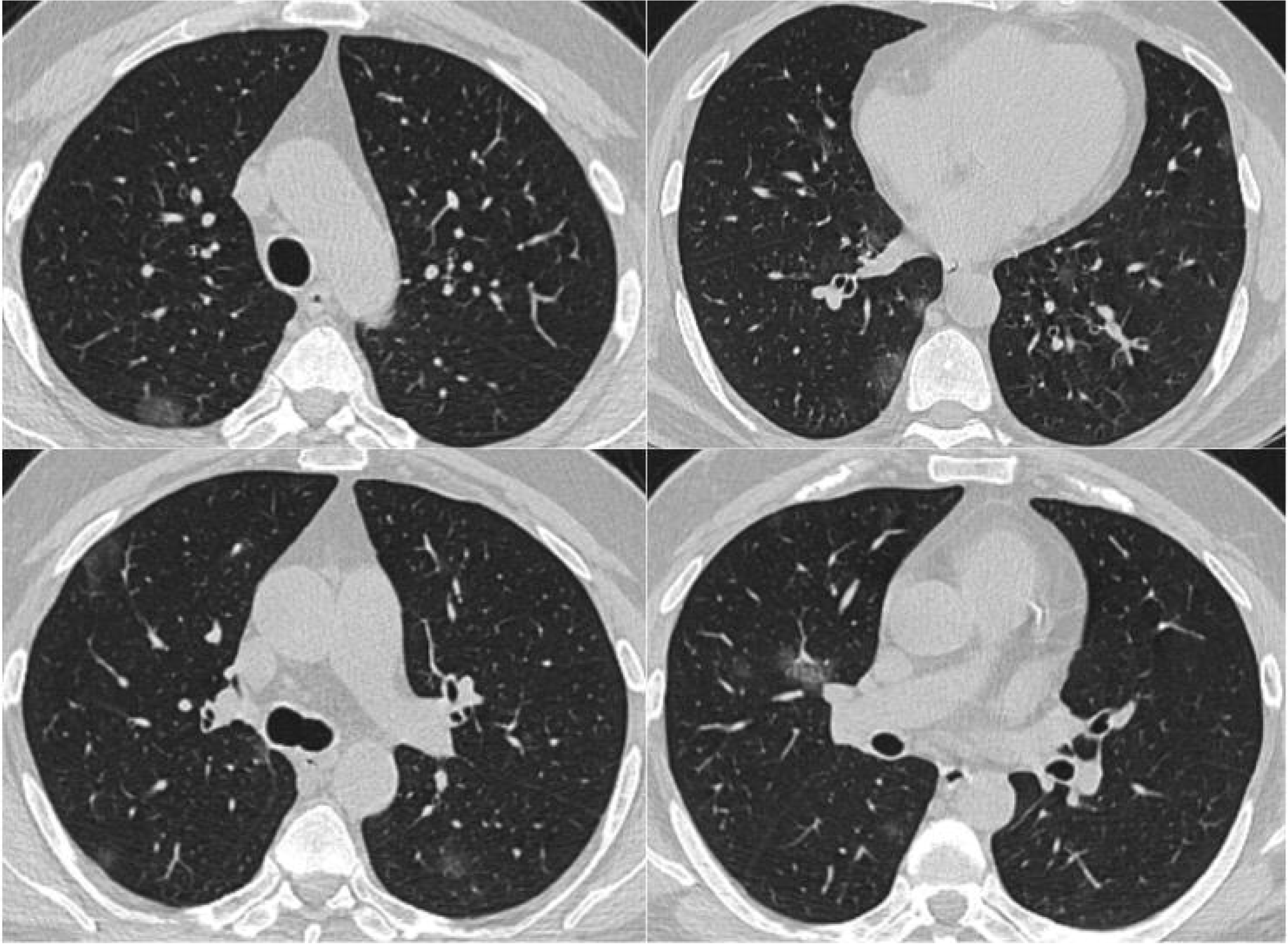
A 59-year-old male patient presented with the complaint of cough. His COVID-19 PCR test was positive. Due to the bilaterally patchy ground glass infiltrates that were seen at thorax HRCT, treatment with hydroxychloroquine plus azithromycin was started.

**Figure 5B.**
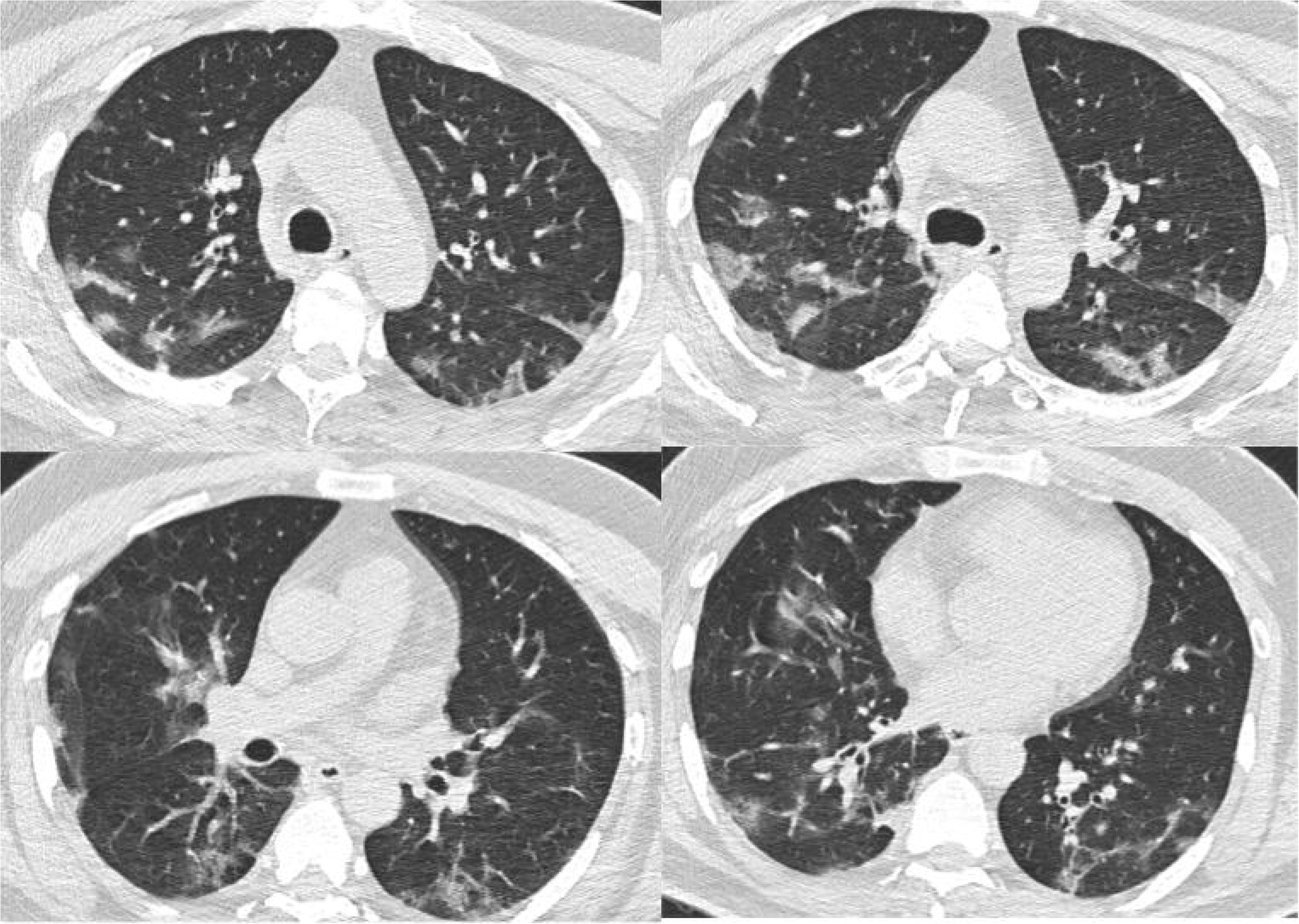
Despite five days of treatment with hydroxychloroquine plus azithromycin, the fever and cough complaints of patient in Figure 5A did not resolve. Since the control CT on the 7th day revealed an increase in the ground-glass infiltrates, treatment with favipiravir plus dexamethasone was started.

**Figure 5C.**
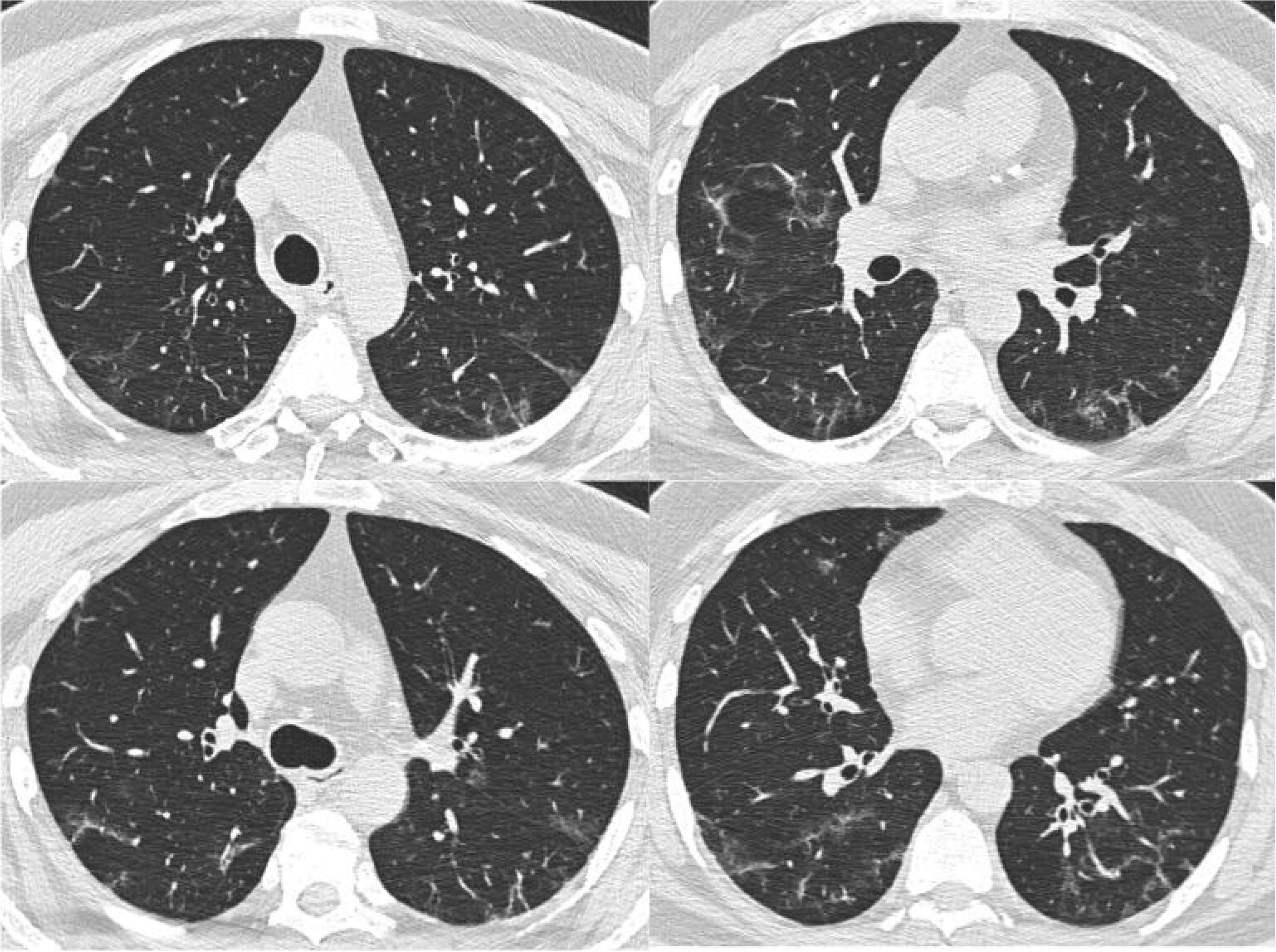
After five days of treatment with favipiravir plus dexamethasone, chest x-ray on the 7^th^ day revealed the regression of lesions and patient’s complaints (cough and fever) have shown a significant improvement.

**Figure 6A.**
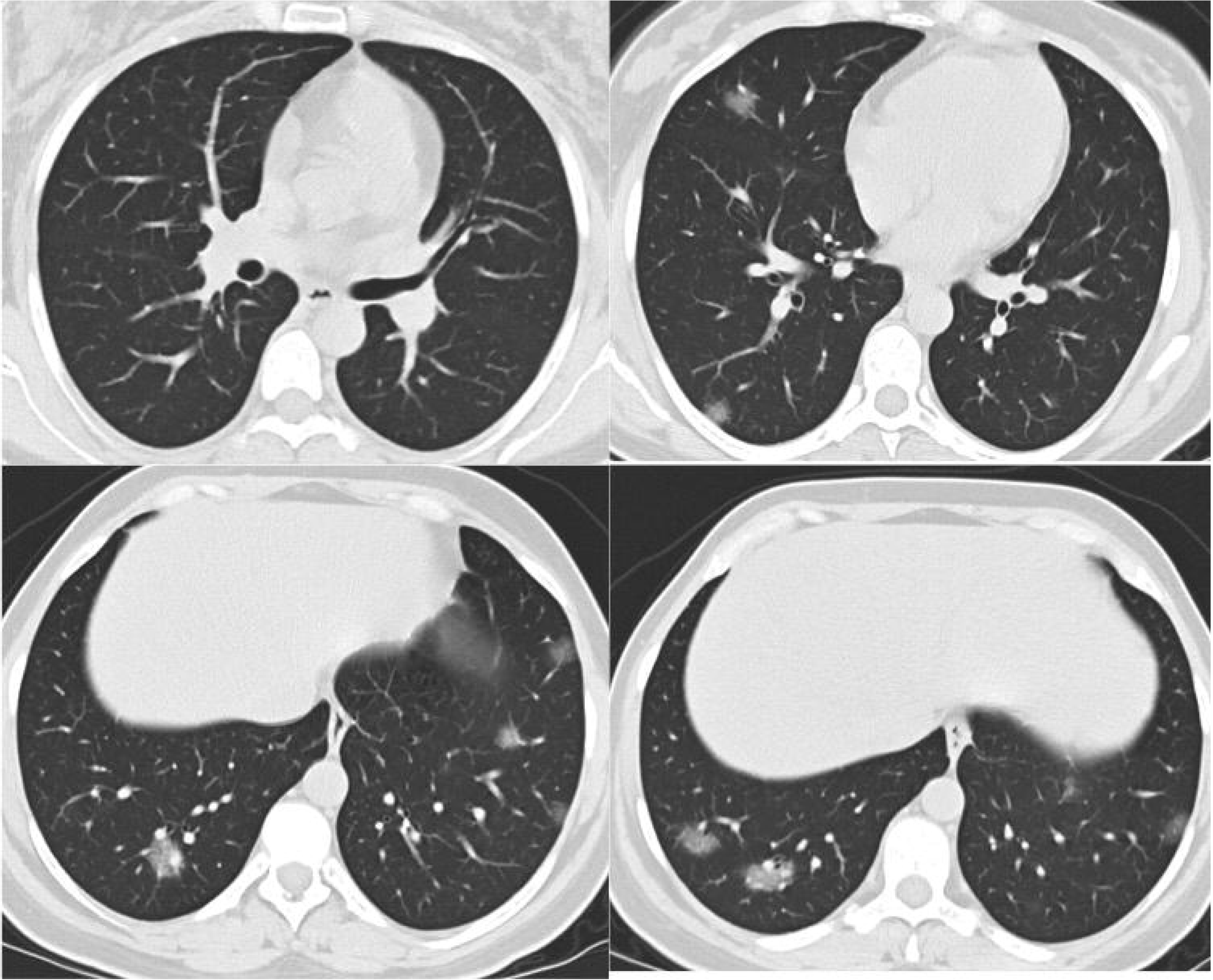
A 32-year-old female patient presented with the complaints of cough, dyspnea and fever. Her COVID-19 PCR test was positive. Due to the bilaterally nodular and ground glass infiltrates that were seen at thorax HRCT, treatment with hydroxychloroquine plus azithromycin was started.

**Figure 6B.**
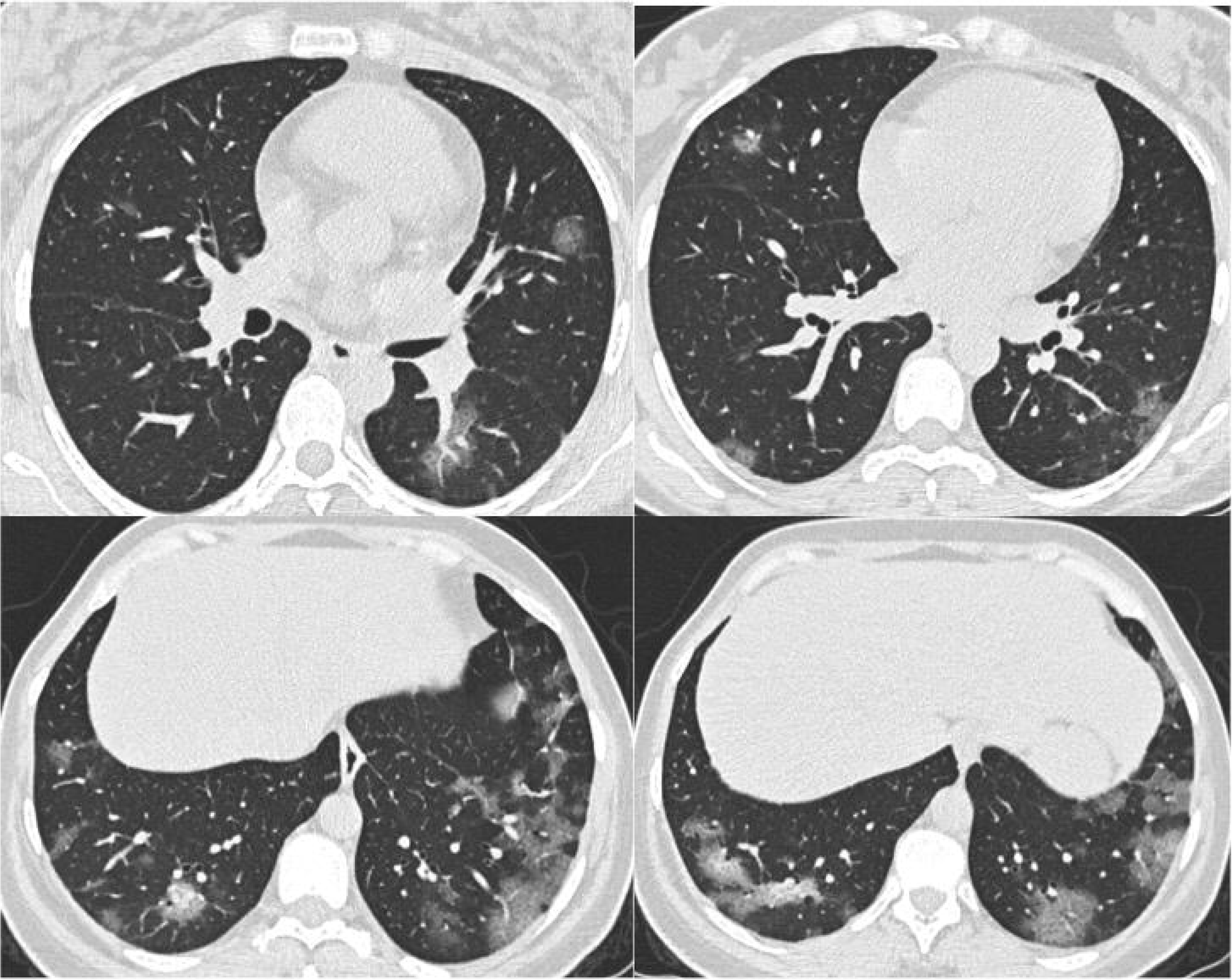
Despite five days of treatment with hydroxychloroquine plus azithromycin, the fever and cough complaints of patient in Figure 6A did not resolve. Since the control CT on the 7th day revealed an increase in the ground-glass infiltrates, treatment with favipiravir plus dexamethasone was started.

**Figure 6C.**
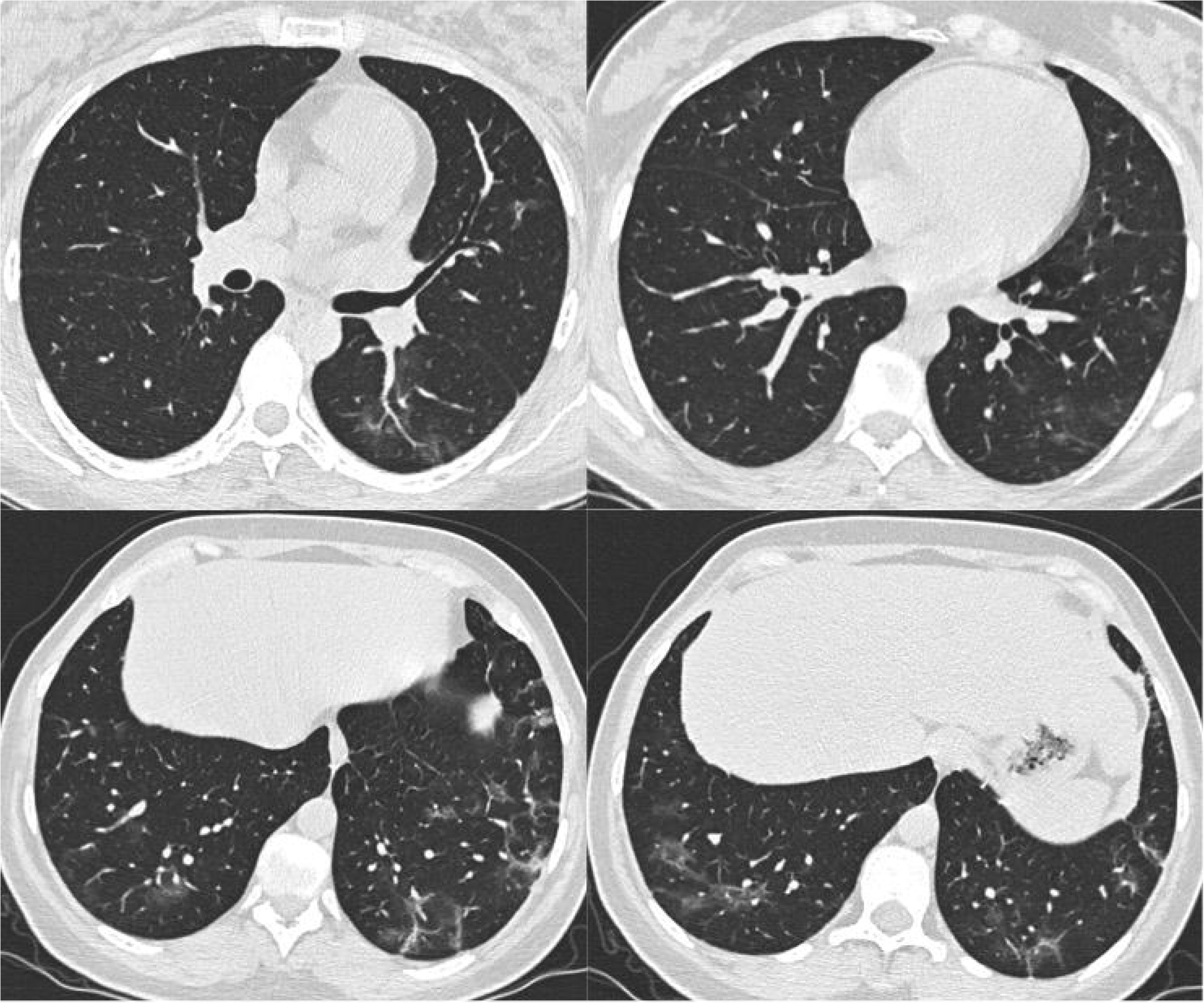
After five days of treatment with favipiravir plus dexamethasone, chest x-ray on the 7^th^ day revealed the regression of lesions and patient’s complaints (cough and fever) have shown a significant improvement.

## Discussion

As of August 16 2020, there is no FDA-approved specific drug for COVID-19. However, an array of the already-existing drug is approved for their other indications, and multiple investigational agents are currently being studied for the treatment of COVID-19 in different clinical trials around the globe (5-14). A large volume of data from randomized controlled trials, observational cohorts, and case series are emerging at a very rapid pace; some in peer-reviewed journals, others as pre-peer-review manuscripts, and, in some cases, in press releases.

On March 9, Yao X. et al. reported that the malaria drug hydroxychloroquine sulfate was effective in killing the SARS-CoV-2 in the laboratory settings (6). In March 2020, the US FDA issued an emergency use authorization (EUA) to allow the emergency use of hydroxychloroquine sulfate supplied from the Strategic National Stockpile (SNS) for the treatment of COVID-19 in certain hospitalized patients. Since then, many national agencies have authorized the use of hydroxychloroquine in COVID-19 patients (7)(8). On June 15 2020, however, the FDA revoked the EUA. Since then, contradictory conclusions have been drawn by many researchers around the world regarding the efficiency of viral clearance and clinical benefit with the regimen of hydroxychloroquine plus azithromycin (7-10). Recent prospective and randomized trials found no evidence of a strong antiviral activity or clinical benefit from the use of hydroxychloroquine plus azithromycin (8-10). Cavalcanti et al. even reported that occurrence of any adverse event was more frequent in patients receiving hydroxychloroquine with azithromycin (7). According to our results, although this combination can be effective in the laboratory conditions, it was not effective on diminishing pulmonary infiltrates of COVID-19 since 65.3% of patients treated with this combination demonstrated a failure of treatment.

Favipiravir is a viral RNA polymerase inhibitor, which acts by its metabolite ribofuranosyl-5’-triphosphate (RTP) that inhibits influenza virus RNA polymerase activity (11). Favipiravir was urgently used and presented good inhibitory effects on the replication of Ebola virus in 2014. Favipiravir is also proven to protect mice from lethal infection with various types of Influenza viruses (12). It is the first approved drug with the potential curative effect on COVID-19 in China, which then played an important role in the prevention and control of SARS-CoV-2 infection (13). Although there have been many registered clinical trials focusing on antiviral drugs for COVID-19, how antivirals may contribute to control the disease progression remains unclear. Q. Cai et al. reported that favipiravir causes an improvement of chest imaging by inhibiting the replication of SARS-CoV-2 (13). They also stated that the relationship between viral clearance and improvement in CT image together helps improvement of the clinical picture of the patient. We observed that, for our patients, favipiravir was the only drug to achieve clinical (especially in fever) and radiological improvement in patients with COVID-19.

Glucocorticoids have been widely used in the syndromes closely related to COVID-19, including Severe Acute Respiratory Syndrome (SARS), Middle East Respiratory Syndrome (MERS), severe Influenza infections and community-acquired pneumonia. Commonly, in all of those severe viral pneumonia cases, the host immune response plays a key role in determining the pathophysiological effects of organ failure (2). The RECOVERY trial has recently demonstrated that dexamethasone reduced the risk for death among seriously ill COVID-19 patients by up to one third (2). However, the drug did not appear to help patients with less serious presentations of COVID-19. The benefit was thought to occur in patients who were being treated more than 7 days after symptoms onset, when inflammatory lung damage likely to constitute the clinical picture (2). Guidelines issued by the U.K. chief medical officers and by the National Institutes of Health (NIH) in the United States have already been updated to recommend the use of glucocorticoids in severe patients hospitalized with COVID-19 (2,4,5,14). In the presented case series, we showed that severe patients who failed to respond to the hydroxychloroquine plus azithromycin combination were successfully treated with favipiravir plus dexamethasone, consistent with the idea of the existence of inflammatory lung environment.

After early May, when the clinicians at our hospital have realized that hydroxychloroquine was not as effective as favipiravir for the recovery of pneumonia patients, all the patients with pneumonia were prescribed with favipiravir instead of hydroxychloroquine. However, creating a randomized controlled group would not be ethical at the time of the pandemic, therefore, we were not able to compare the effects of drug by the matched randomized group. Instead, we tried to compare the before after results of two different patient populations treated at different time intervals. In our perspective, it is important to distinguish COVID-19 patients who developed pneumonia and adjust a new treatment for them according to clinical severity of each patient.

The most important point to be illuminated in patients with COVID 19 pneumonia; is the question of whether lesions in the lung are infective(pneumonia) or inflammatory(pneumonitis). Radiology and pathology examinations of patients with COVID-19 revealed inflammatory reactions in the lung that resembled what is observed in hypersensitivity pneumonitis rather than in other viral pneumonia (15,16). Tian S et al. reported histopathology data of COVID-19 pneumonia deriving from the accidental cases of two patients who underwent lobectomies for lung cancer (16). Pathologic findings were edema, proteinaceous exudate, vascular congestion, inflammatory clusters with multinucleated giant cells, interstitial fibroblastic proliferation, and reactive hyperplasia of pneumocytes. Since patients had no symptoms at the time of surgery, the authors conclude that these features could represent an earlier phase of COVID-19 pneumonia. Instead, in the advanced stages of the disease, the main pathological findings are diffuse alveolar damage with lymphocytic infiltrate, small thrombotic vessels, and foci of alveolar hemorrhage (16-18). Supportively, Polak et al. mentioned the three stages of COVID-19 being the viral replication, inflammation with coagulopathy and fibrosis phases, all occur either simultaneously or consecutively (19). Potus et al. also stated that endothelial dysfunction and microthrombosis associated with SARS-CoV-2 infection is strikingly similar to what is seen in pulmonary vascular diseases. They also stated that for this reason, patients with preexisting pulmonary vascular disorders with altered endothelial cell metabolism are at great risk of mortality (20). Thus, awareness regarding the precise nature of the immunological reaction on the entire vasculature and the lung tissue in response to viral infection is essential to initiate timely and targeted supportive therapy.

Supportively, we concluded that; SARS-CoV-2 first starts as an infection of the nasopharynx and progresses as the inflammation of the lung parenchyma, depending on the degree of the response that is shown by the host immunity. We also think that pulmonary radiological findings that appear as opacities are also caused by the inflammatory reaction against the virus. The response to the anti-inflammatory therapy that is proven by radiological and clinical improvement, as well as published pathological data all support this finding. However, one limitation of this study includes a lack of pathological investigation of patients due to the inability to take a biopsy during clinical practice due to the circumstances during the pandemic.

Segmental small pulmonary vessel enlargement on chest CT has been stated as a unique feature associated with COVID-19, different from other viral or bacterial pneumonia findings on chest CT (21). Lang et. al also stated that medium to small vessel dilatations are not confined to areas of the diseased lung, and often involve subpleural distal vessels, suggesting a diffuse vascular process (22). One theory behind those findings is the possibility of COVID-19-induced secondary pulmonary vasculitis that is responsible for vascular changes and abundance of pulmonary parenchymal ground-glass opacification (21, 22). The vascular dilatations seen in ground glass-opacities of COVID-19 can be a good indication to differentiate COVID-19 from other etiologies of ground-glass opacities, as seen in our cases (Figure 7 and 8). Those vascular dilatations might indicate that COVID-19 starts as vasculitis and progresses to the perivascular inflammation. It is not clear if it is an immune-complex vasculitis or not, and further pathological examinations must be done to have a clear result.

**Figure 7.**
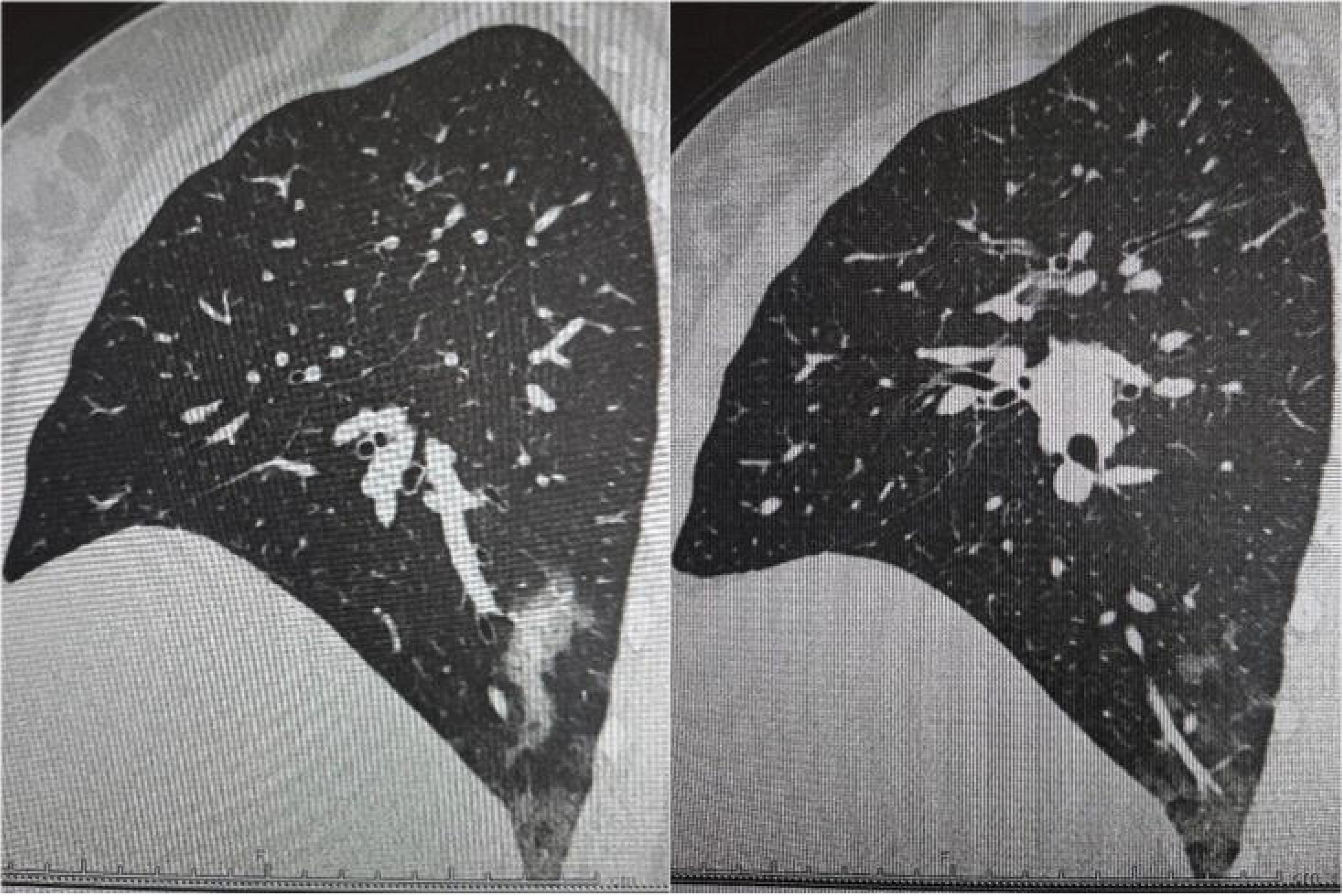
The vascular enlargement and perivascular ground glass opacities (GGOs) in a 32 years-old female patient with COVID-19 pneumonia.

**Figure 8.**
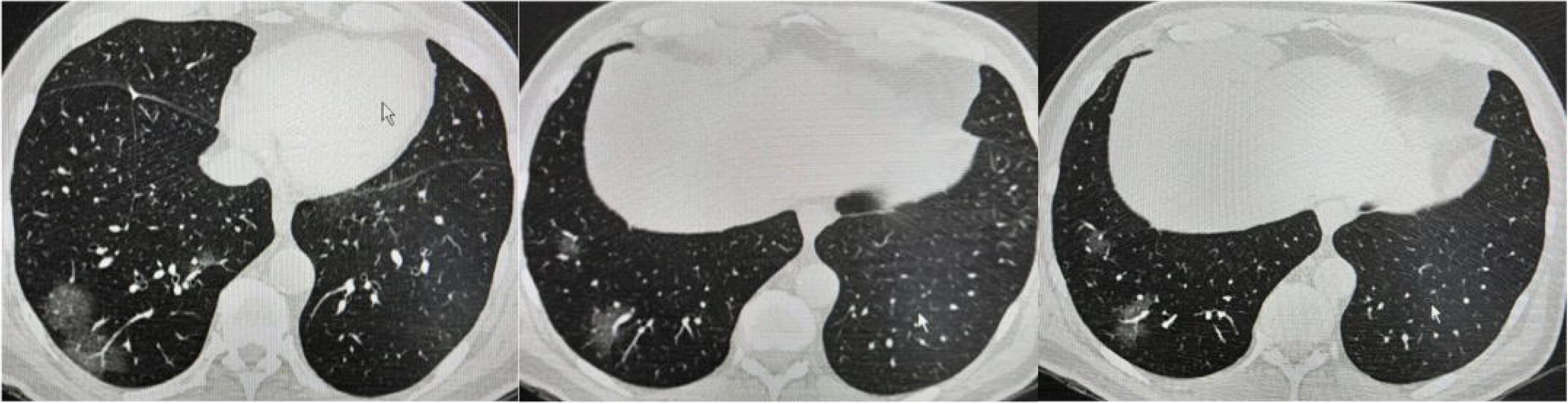
The vascular enlargement and perivascular ground glass opacities (GGOs) in a 70 years-old male patient with COVID-19 pneumonia.

This study has some limitations. Since this study was not designed to compare the two groups at the same time but rather to compare the outcomes of two different groups of patients treated at different times retrospectively, the study lacks the features of a randomized control trial. Future studies with matched control groups and patients with other appropriate pathologies, including bacterial or other types of viral pneumonia would be helpful to explain the findings and clarify the extent of the vascular findings. Well-designed large randomized clinical trials are also necessary to identify effective treatments based on a high level of evidence. Further imaging and pathologic studies are also required to investigate the diffuse pulmonary vascular dysregulations that are caused by unclear mechanisms.

In conclusion, we suggest that the SARS-CoV-2 causes an infectious disease that progresses to inflammatory lung disease. The pulmonary infiltrates seem to be not infective; therefore, the characteristic of the disease should be described as COVID-19 pneumonitis instead of pneumonia. We concluded that, although the hydroxychloroquine plus azithromycin can be used effectively in the infected people without pneumonia, there is no clinical benefit from the use of such a combination in infected patients who developed pneumonia. The favipiravir plus dexamethasone seems to be the only drug combination to achieve the improvement of radiological presentation and clinical symptoms in severe COVID-19 patients.

## Data Availability

The research article data that used to support the findings of this study is available from the corresponding author upon request.

## Acknowledgments

None

